# Short-term survival benefit associated with neonatal clinical trial participation: An observational cohort study in The Gambia

**DOI:** 10.64898/2026.08.04.26359594

**Authors:** H Brotherton, A Gai, G Walker, Y Njie, S Kapoor, A Hough, M Bittaye, U Okomo, S Cousens, A Roca, JE Lawn

## Abstract

**Background:** Trial participation effect, defined as a change in clinical outcomes associated with trial enrolment regardless of allocation, is understudied in neonatal trials in low- and middle-income countries (LMICs), despite its importance for trial design, interpretation, and research ethics. This study aimed to quantify the trial participation effect and explore potential ways by which research participation may influence neonatal survival.

**Methods:** This observational cohort study included neonates weighing <2Kg and aged <24h who were admitted to a Gambian referral hospital and either enrolled in a clinical trial comparing early versus later KMC (eKMC trial;2018-2020) or not enrolled due to operational constraints and hence received standard, non-research care. All infants were prospectively followed until in-patient discharge or death. The eKMC trial previously found no important effect of early KMC on all-cause neonatal mortality. For this analysis, in-patient mortality rates were compared using a generalised linear model, adjusting for baseline differences in participant characteristics. Prospectively collected data on small and sick newborn care readiness and delivery during the trial period were used to explore how trial participation may have influenced survival.

**Results:** A total of 545 neonates were included: 279 enrolled in the trial and 266 not enrolled, predominantly due to the absence of an available caregiver. Baseline characteristics were similar between groups, although differences were seen in twin status, place of birth, and age at admission. Trial participation was associated with an absolute reduction in inpatient mortality of 6.3% (22.6% (63/279) among enrolled versus 28.9% (77/266) among non-enrolled) and a relative reduction of 29% (aRR 0.71, 95% CI 0.53–0.96). This association varied by season, with no evidence of benefit during the dry season (aRR 0.97, 95% CI 0.60-1.58), but a 40% reduction in adjusted mortality risk during the rainy season (aRR 0.60, 95% CI 0.41-0.87)(Interaction test: *p*=0.086). Trial participants had access to laboratory diagnostics and received more intensive clinical monitoring, including higher staffing ratios, continuous pulse oximetry, structured education of carers on neonatal danger signs, and enhanced scrutiny of clinical management compared to neonates receiving routine care.

**Conclusion:** Trial participation was associated with a substantial reduction in inpatient mortality, suggesting that participation effects should be considered when designing, interpreting, and reporting neonatal clinical trials in LMIC settings. The association was evident only during the rainy season. The participation effect may have been mediated by increased clinical oversight and monitoring, additional nursing support, and access to diagnostic investigations, all of which should be prioritised within routine care to accelerate progress towards SDG neonatal survival targets.

**Key messages:** **What is already known on this topic**

- It is widely perceived that patients in low- and middle-income countries (LMICs) may benefit from participation in clinical trials because of additional resources and access to otherwise-unavailable health services.
- The magnitude of the trial participation effect on neonatal survival in LMICs has not previously been quantified

**What this study adds**

- *This is the first study from an LMIC to quantify an association between participation in an individually randomised neonatal clinical trial and reduced inpatient mortality.*
- Preterm/low birth weight (LBW) newborns enrolled in the eKMC trial had 29% lower adjusted relative risk of inpatient mortality compared to clinically comparable, non-enrolled infants receiving routine care at the same neonatal unit
- The association between trial participation and reduced mortality was observed only during the rainy season (July-October), when mortality rates were highest, suggesting that participation effects may be most important in higher-mortality contexts.
- Trial participants received enhanced clinical monitoring and higher-quality small and sick newborn care (SSNC), including continuous pulse oximetry, higher nursing availability, rigorous safety oversight, and access to laboratory diagnostics, especially blood cultures.
These findings demonstrate that preterm/LBW newborns can safely participate in clinical trials in resource-limited settings when essential components of SSNC are in place.
- The study identified key elements of SSNC that should be prioritised within health-system strengthening efforts to accelerate progress towards SDG 3.2.

**How this study might affect research, practice or policy**

- Research: These findings provide the first quantifiable estimate of a neonatal trial participation effect in a low-resource, high-mortality hospital setting. This information could improve the accuracy of sample size calculations and interpretation of trial findings, reducing the risk of underpowered studies and research inefficiency. The findings also highlight the ethical importance of measuring and reporting trial participation effects, particularly in settings where research activities may strengthen the quality of routine newborn care and where effects may be greatest during periods of high mortality risk.
- Clinical practice: Key components of SSNC that differed between trial participants and non-participants, included continuous monitoring, higher nursing availability, regular safety oversight, and access to microbiological diagnostics. Strengthening these elements within routine care could improve outcomes for preterm/LBW neonates regardless of research participation and especially in highest mortality settings.
- Policy: Investments in neonatal monitoring capacity, nursing, safety and clinical governance, and microbiology services should be prioritised within national newborn health strategies to accelerate progress towards SDG3.2. The greater impact observed during the high-mortality rainy season suggests that such investments may yield the largest survival gains in the highest burden, vulnerable global contexts.

## INTRODUCTION

Global child mortality has declined substantially over the past decade, yet reductions in neonatal mortality have been far slower [1]. Worldwide, 2.3 million newborns still die each year, with 98% of deaths occurring in low and middle-income countries (LMICs), and recent estimates indicate stalled progress [1]. Sustainable Development Goal (SDG) 3.2 aims to reduce neonatal mortality rates to <12 per 1000 live births by 2030. However, with less than 4 years remaining and 64 countries currently off-track [1], accelerating neonatal survival in LMICs is a public health priority. Gains are especially critical for neonates born preterm or with low birth weight (LBW), who experience the highest morbidity and mortality of all children across all income-settings.

Participation in clinical trials is often perceived to confer individual benefit through enhanced clinical care compared with routine services, independent of any intervention effect. This phenomenon, known as the *trial participation effect* [2], has been demonstrated in several high-income country (HIC) populations, including adult oncology [2, 3], stroke research [4], and women’s health [5], with limited evidence in paediatric populations [3, 6] and very little in neonates [7-9]. The few neonatal studies that have examined trial participation effects were all conducted in HIC hospitals, where health-system capacity and baseline mortality rates differ markedly from low-resource LMIC settings. As a result, the existence and magnitude of any neonatal trial participation benefit for LMICs is currently unknown.

Understanding the neonatal trial participation effect is important for methodological, ethical and clinical reasons. Quantifying any survival difference could improve the accuracy of sample size calculations for trials evaluating neonatal mortality, supporting trials that are adequately powered and reducing research inefficiency. This is particularly relevant given the under-representation of neonatal research in global funding portfolios [10]. In LMICs, obtaining access to improved care is a key motivator for families considering research participation [11, 12], and a clear understanding of potential benefits and mechanisms is essential for transparent and informed consent. Furthermore, identifying context-specific pathways through which research involvement may influence neonatal outcomes could highlight priorities for prioritising and strengthening the quality of SSNC in low-resource LMIC health-facilities.

This study addresses this knowledge gap by comparing in-hospital mortality rates among preterm/LBW neonates who participated in a clinical trial with those of clinically comparable neonates receiving usual SSNC in a low-resource Gambian health-facility. A secondary aim was to prospectively examine differences in SSNC access and provision between the two groups and identify potential factors associated with survival differences. This study is reported in accordance with STROBE guidelines [13].

## METHODS

### Design

This observational cohort study was embedded in a randomised controlled trial investigating the survival and clinical effects of early Kangaroo Mother Care (KMC) for unstable small newborns (eKMC trial: clinicaltrials.gov ref:NCT03555981). No evidence for an intervention effect was identified by the eKMC trial, although power to detect a difference in the primary outcome (all-cause mortality by chronological day 28) was low [14]. Additionally, the daily duration of KMC delivered to eKMC trial participants in both the control and intervention groups [14] was less than the threshold needed for a KMC-related mortality effect (8h/day) [15, 16], hence an intervention effect was not expected and all the trial cohort were included in this ancillary analysis.

### Setting

The eKMC trial was conducted at Edward Francis Small Teaching Hospital (EFSTH), the only teaching hospital and neonatal referral centre in The Gambia, West Africa. The neonatal mortality rate (NMR) in The Gambia was estimated to be 26 deaths per 1000 livebirths at trial onset.[17] EFSTH’s neonatal unit admits approximately 1400 neonates/year with seasonal variation in birth rates, admission rates and in-patient mortality rates, with higher observed rates during the rainy season (July-October) [18]. As the national referral centre, EFSTH admits neonates born at EFSTH maternity unit (in-born) and other primary or secondary government health facilities and private clinics (out-born). During the last decade, neonatal case fatality rates (CFR) at EFSTH ranged from 33% (2010) to 39% (2013) for all admitted neonates, with 48% CFR for neonates weighing <2kg [18]. An estimated 12% of Gambian neonates are born premature, defined as <37 weeks gestation, with 17% LBW, [1] yet complications of being born preterm or LBW account for 39% of all neonatal deaths at EFSTH [18].

WHO level 2+ SSNC [19] was available at EFSTH during the study period and delivered according to a standardised guideline aligned with WHO recommendations for preterm/LBW neonates [20] and co-developed by the eKMC team and EFSTH clinical staff. Bubble CPAP was the highest level of respiratory support, and oxygen was supplied via concentrators that typically served multiple neonates simultaneously, with no flow-splitters for titration. Intravenous (IV) maintenance fluids were manually prepared and administered through burettes, with the single fluid pump prioritised for blood transfusions. Essential medications including vitamin K, ampicillin, gentamicin, flucloxacillin, ciprofloxacin and aminophylline were routinely available, while caffeine citrate and cephalosporins were available intermittently. As part of the eKMC trial, additional supplies of caffeine citrate, vitamin K, meropenem and additional consumables (e.g., cannulas, syringes) were provided for all admitted neonates, regardless of trial enrolment.

Kangaroo mother care (KMC) was introduced at EFSTH neonatal unit in 2017 as standard care for preterm/LBW neonates, either intermittently at the cot-side or as continuous KMC in an 8-bed KMC unit. Neonates were transferred from the neonatal unit to the KMC unit once stable on full enteral feeds and no longer requiring CPAP or oxygen. eKMC trial participants received KMC according to standard care (control) or within 24h of admission (intervention), alongside routine supportive care. To facilitate early KMC, trial participants were managed in a designated area of the neonatal unit, with beds for mothers/carers in the intervention group and incubators/radiant heaters for the control group. Similar to non-enrolled neonates, eKMC participants were transferred to the KMC unit once stable off oxygen and IV fluids [15].

### Participants

All neonates <2 kg admitted to the site and screened for recruitment into the eKMC trial between May 2018 and March 2020 were potentially eligible for this ancillary study. To ensure comparability between groups, the original eKMC clinical eligibility criteria [21] were applied post-hoc to the full cohort of screened neonates to identify neonates who met clinical eligibility criteria but were not enrolled due to operational or consent-related reasons. Eligible newborns were singleton or twins <2kg and <24 hours old at admission, who were mild-moderately unstable and without seizures, severe jaundice or major congenital malformations [21]. Neonates with missing outcome data (discharge or death) were excluded (n=16, 2.8% of those meeting eligibility criteria).

### Data collection

During eKMC trial screening procedures, parents or carers of neonates <2 kg were informed about the trial before socio-demographic and clinical data were collected. Trained research staff used electronic case report forms on RedCAP to record screening data. Admission age was confirmed using time of birth data from maternal hand-held records or referral letters; weight was measured using calibrated digital scales. Research clinicians supervised examinations for congenital malformations and severe jaundice. Axillary temperature was measured using digital thermometers (three measurements with mean value calculated). Heart rate and oxygen saturation were assessed every minute for 10 minutes using a Nonin 2500A pulse oximeter, with manual counting of respiratory rate; mean values were automatically calculated. All assessments followed standardised, protocol-drive procedures with regular training and internal standardisation [21]. The highest level of respiratory support during the screening period, lasting between 10 minutes to 3 hours, was recorded by direct observation. A validated mortality risk score (NMR-2000 score) was calculated post-hoc using admission weight, mean oxygen saturation and level of respiratory support [22]. Data on post-natal gestational age assessment, KMC daily duration, and length of stay were not available for non-enrolled neonates and therefore were excluded from analyses. In-hospital mortality for all screened neonates was recorded prospectively using direct observation, NNU ward register, and death certificates as part of eKMC trial procedures.

### Evaluation of small and sick newborn care

Health-facility service readiness assessments were conducted every 4-6 months during eKMC trial recruitment period (May 2018 to March 2020) to compare care available to enrolled and non-enrolled neonates. A standardised survey was completed by trial personnel using direct observations and interviews with senior nursing and medical staff. Data included availability and functionality of equipment, consumables, medications, investigations, and staffing levels. These assessments were supplemented by eKMC standard operating procedures describing clinical monitoring and safety oversight. Staff-to-patient ratios for the usual-care group were calculated using prospective admission audit data, with ratios for eKMC trial group calculated according to monthly recruitment data.

### Statistical analysis

The primary exposure of interest was enrolment within the eKMC trial. Baseline socio-demographic and clinical characteristics were compared between groups using the Wilcoxon-Mann-Whitney test for continuous variables and Chi squared test for categorical variables. The primary outcome was the in-patient case fatality rate (CFR), defined as the proportion of deaths among all neonates meeting inclusion criteria. CFR was compared between exposure groups (enrolled versus not enrolled) using a generalised linear model (binomial distribution, log link), adjusting for baseline variables that differed significantly between groups and are evidence-based predictors of neonatal mortality (e.g., twin status, out-born delivery with ex-utero transfer). Season was examined as a potential effect modifier given marked seasonal variation in neonatal admissions and mortality at the study site [18], by fitting a log-binomial generalised linear model with an interaction term between research participation and season, in-order to present season-stratified adjusted estimates. Missing data were assumed to be missing completely at random, with complete case analysis used. Analyses were conducted using STATA version 18.

### Ethical approval

Ethical approval for this ancillary study was granted by the Gambian Government/MRC Gambia at LSHTM Joint Ethics Committee (reference number 1643) and the LSHTM Interventional Ethics committee (reference number 16189) as part of eKMC trial approvals. Caregivers of eKMC trial participants provided written informed consent for data collection. Caregivers of non-enrolled non-enrolled participants provided verbal assent for collection of clinical and basic socio-demographic data as part of eKMC trial screening process. Additional consent for re-use of data was not required as this secondary analysis used de-identified, routinely collected outcome data.

## RESULTS

Of 1107 screened neonates, 545 met clinical eligibility criteria and had outcome data, of which 279 were enrolled to the eKMC trial and 266 received usual care (Figure 1). The most common reason for non-enrolment was caregiver unavailability during first 24h of hospital admission (159/266, 59.8%), followed by unavailability of a study bed (69/266, 25.9%) (Supplementary Table 1).

**Figure 1.**
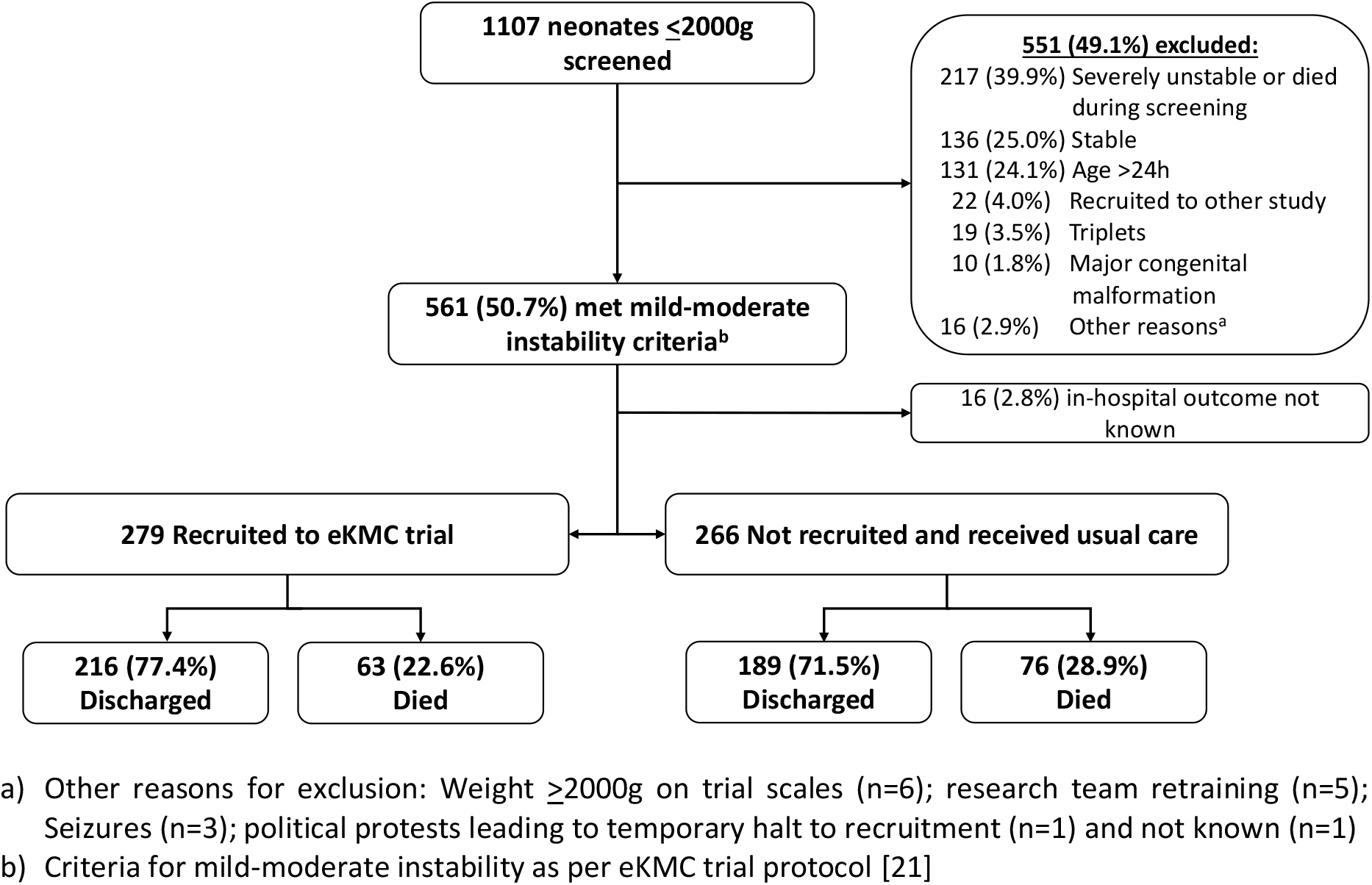
Overview of neonatal participants and outcomes included in a secondary analysis to estimate trial participation effect

### Baseline neonatal characteristics

Neonates enrolled into the eKMC trial were older at admission (2.3h versus 1.2h old, *p*=0.003), with a higher proportion of twins (31% versus 23%, *p*=0.038) and less likely to be in-born (47% versus 61%, *p*=0.001) compared to non-enrolled neonates. All other baseline clinical features were similar between groups, including admission temperature, NMR-2000 mortality risk score, sex and admission weight (Table 1).

**Table 1.** Baseline characteristics of neonates <2 Kg who were enrolled versus not enrolled to a small and sick newborn clinical trial.

|  | Enrolled<br>N= 279 | Usual care<sup>a</sup><br>N= 266 | p value |
| --- | --- | --- | --- |
| Male sex, N° (%) | 118 (42) | 117 (44) | 0.547 |
| Weight (g), median (IQR) | 1450 (1192 – 1650) | 1500 (1200 – 1742) | 0.089 |
| Weight distribution, N° (%) |  |  | 0.708 |
| <1200g | 71 (25) | 64 (24) |  |
| ≥1200g | 208 (75) | 202 (76) |  |
| Twin, N° (%) | 86 (31) | 61 (23) | 0.038 |
| Admission age (h), median (IQR) | 2.3<sup>b</sup> (0.78 – 5.25) | 1.2<sup>c</sup> (0.63 – 3.28) | 0.003 |
| Admitted during night shift,<sup>d</sup> N° (%) | 119<sup>e</sup> (43) | 115<sup>f</sup> (44) | 0.739 |
| Admitted during rainy season,<sup>g</sup> N° (%) | 136 (49) | 140 (53) | 0.364 |
| In-born (EFSTH),<sup>h</sup> N° (%) | 132 (47) | 163 (61) | 0.001 |
| Axillary temperature (°C), median (IQR) | 36.1<sup>i</sup> (35.5 – 36.8) | 36.2<sup>j</sup> (35.5 – 36.9) | 0.223 |
| Respiratory rate (CPM), median (IQR) | 58<sup>k</sup> (49 – 70) | 58<sup>l</sup> (49 – 68) | 0.848 |
| Heart rate (BPM), median (IQR) | 139<sup>m</sup> (128 – 153) | 141<sup>n</sup> (132 – 156) | 0.105 |
| SpO<sub>2</sub> (%), median (IQR) | 98<sup>o</sup> (95 – 99) | 97<sup>n</sup> (95 – 98) | 0.174 |
| SpO<sub>2</sub> <88%, N° (%) | 16/271 (6) | 14/235 (6) | 0.980 |
| NMR-2000 score<sup>p</sup>, median (IQR) | 17.6<sup>q</sup> (14.8 – 19.6) | 18.4<sup>r</sup> (15.1 – 20.3) | 0.053 |

BPM = beats per minute; CPM = cycles per minute; EFSTH = Edward Francis Small Teaching Hospital; IQR = Interquartile range.
a) Eligible but not recruited due to lack of study bed, absence of caregiver within 24h of admission or consent declined; b) n = 268; c) n = 246; d) Night shift defined as 20:00 – 07:59; e) n = 278; f) n=260; g) Rainy season from June to October; h) Data on place of birth not collected during screening, but place of referral assumed to be place of birth due to age <24h at admission; i) n = 266; j) n = 243; k) n = 259; l) n= 255; m) n = 207; n) n = 191; o) n= 206; p) NMR-2000 score consists of birth weight, admission SpO<sub>2</sub> and highest level of respiratory support. Low risk of mortality is indicated by NMR2000 score >16 [22]; q) n = 210; r) n = 181;

### Health service readiness for small and sick newborn care

Enrolled neonates received enhanced clinical monitoring and oversight of SSNC compared to neonates receiving usual care (Table 2). The enhanced clinical monitoring strategies linked to research participation were intended to ensure participant safety and included: continuous pulse oximetry until neonates were stable off oxygen; more frequent monitoring of vital signs, especially during the first 24h of enrolment; increased nursing and clinician staff to patient ratios; and structured education of caregivers about danger signs. Medical oversight of SSNC was protocolised for trial participants with daily scrutiny of management plans by trained clinicians to ensure provision of standardised, WHO-compliant clinical care, active surveillance for adverse events and scrutiny of the SSNC delivered to all severely unwell and deceased neonates by two Consultant Paediatricians, trial sponsor, and UK/Gambian ethical committees. Neonates receiving routine care did not have access to any formal morbidity or mortality audit process. Trial participants had free access to high quality MRC Unit The Gambia laboratory diagnostics such as blood culture and basic biochemical tests (serum bilirubin, urea and electrolytes), which were not available to non-enrolled neonates at EFSTH (Table 2). Health service readiness for SSNC did not vary during the trial period for either group, with no seasonal variation in access to clinical monitoring, additional staff, or diagnostics.

**Table 2.**
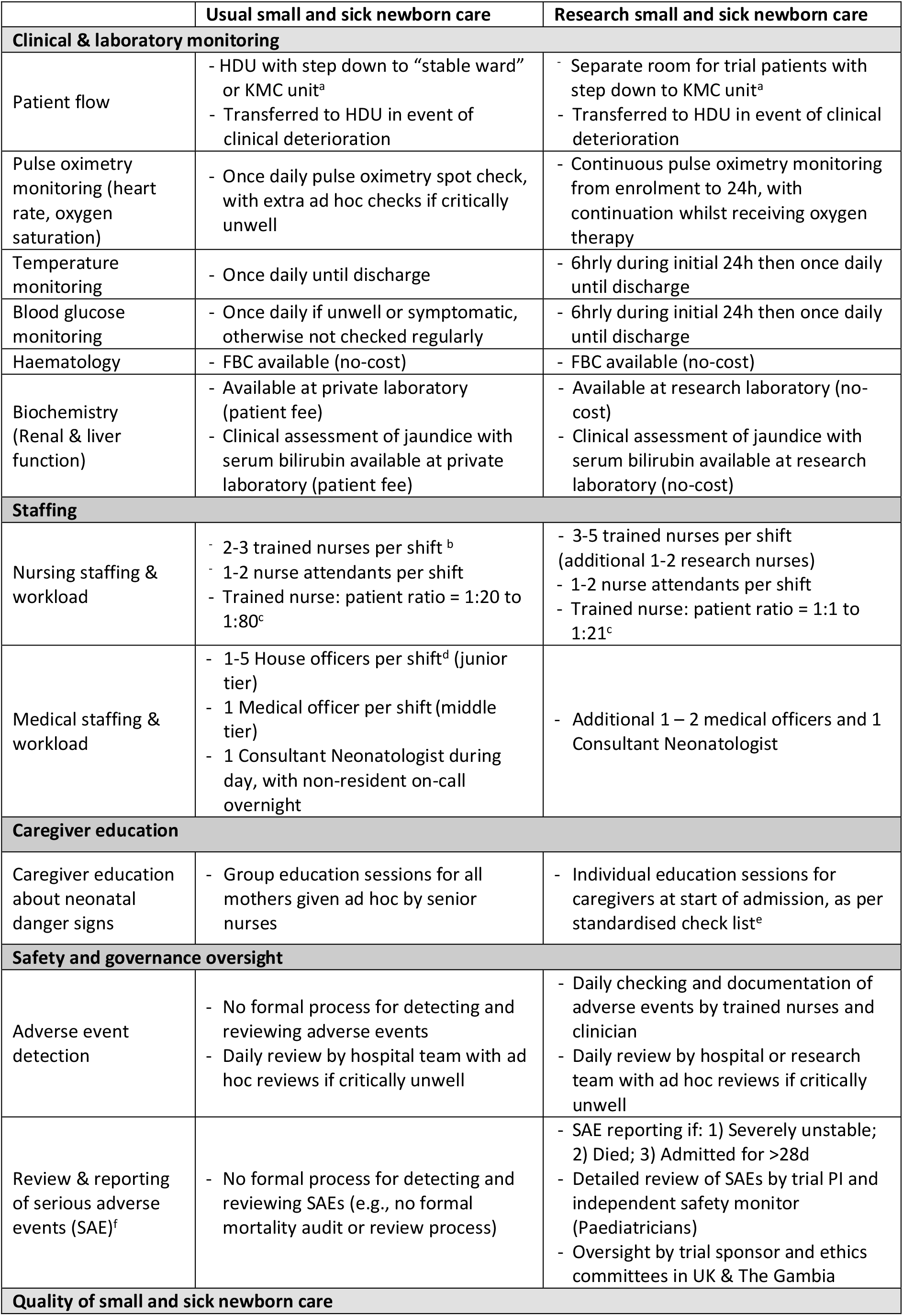
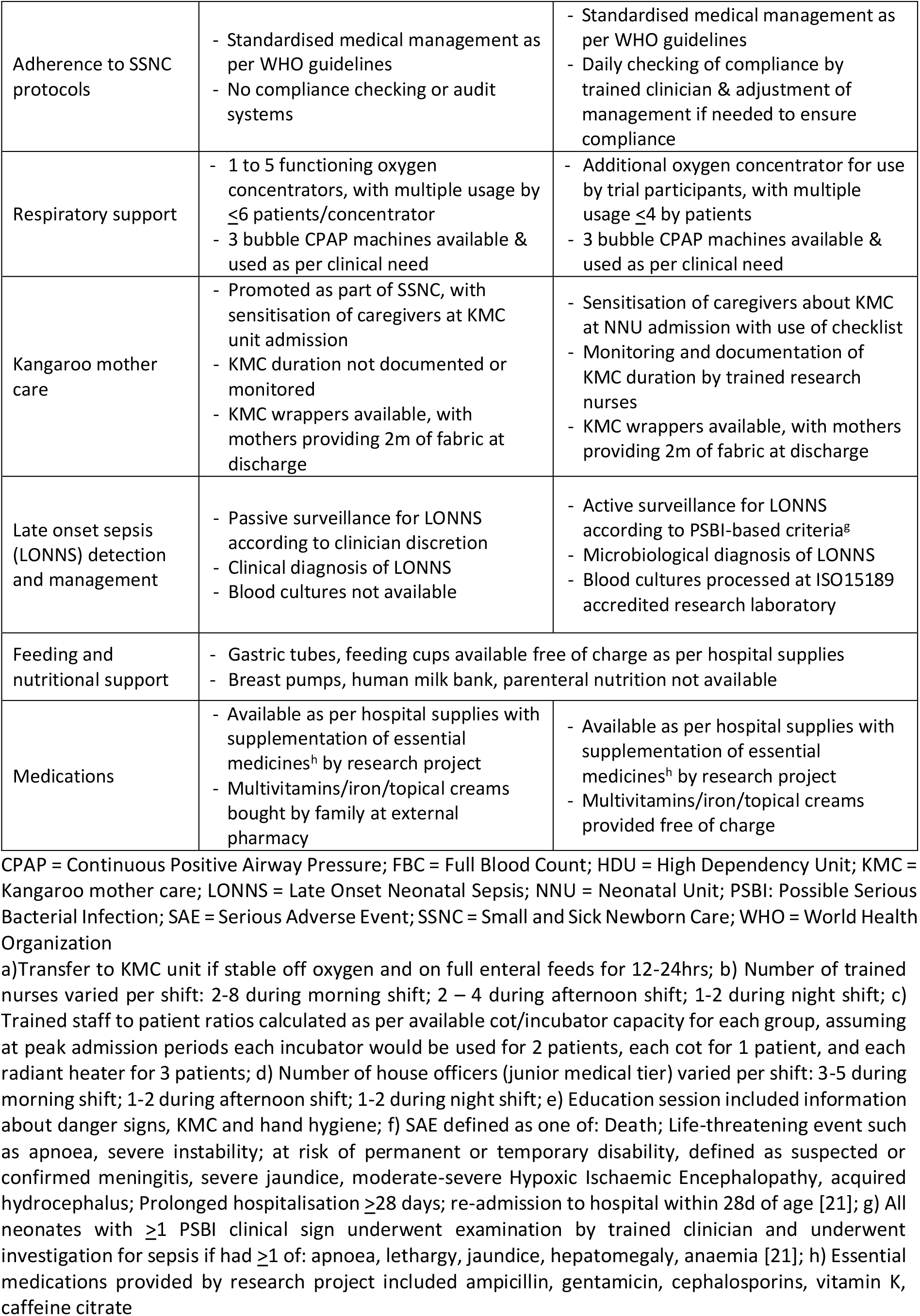
Comparison of clinical care available to small vulnerable neonates as part of usual versus research care.

### Effect of research participation on inpatient neonatal case fatality rates

Overall, neonates recruited to eKMC trial had a 22% reduced relative risk of inpatient mortality (22.6%, 63/279) compared to clinically comparable newborns not enrolled (28.9%, 77/266), with 29% reduced relative risk after adjustment for admission age, twin and in-born status (aRR 0.71, 95% CI 0.53 – 0.96, *p* = 0.024). This absolute risk difference of 6.3% is equivalent to 6 fewer deaths per 100 neonatal admissions. The inpatient CFR for usual-care neonates varied according to season of admission, with higher rates during rainy season (52/140, 37.1%) versus the dry season (25/126, 19.8%). Research participation was not associated with inpatient neonatal mortality during the dry season (aRR 0.97, 95% CI 0.60 to 1.58), but was associated with 40% reduced relative risk of mortality during the rainy season (aRR 0.60, 95% CI 0.41 to 0.87) (Table 3). Evidence for an interaction between research participation and season of admission was weak (*p*=0.086), indicating independent associations.

**Table 3.** Association between research participation and inpatient neonatal mortality overall and stratified by season.

| Population | In-patient mortality |  | Unadjusted analysis |  | Adjusted analysis<sup>a</sup> |  |
| --- | --- | --- | --- | --- | --- | --- |
|  | Research neonates | Usual care neonates | Crude RR (95% CI) | P value | Adjusted RR (95% CI) | P value |
| Total cohort (n=545) | 63/279 (22.6) | 77/266 (28.9) | 0.78 (0.59-1.04) | 0.09 | 0.71 (0.53-0.96) | 0.024 |
| Rainy season (n=261) | 33/136 (24.3) | 52/140 (37.1) | 0.65 (0.45-0.94) | 0.023 | 0.60 (0.41-0.87) | 0.007 |
| Dry season (n=251) | 30/143 (21.0) | 25/126 (19.8) | 1.06 (0.66-1.70) | 0.818 | 0.97 (0.60-1.58) | 0.908 |

Nº = Number; RR = Risk ratio
a) Adjusted risk ratios (RRs) are from log-binomial generalised linear models adjusted for twin status, place of birth, and age at admission

## DISCUSSION

Preterm and LBW neonates enrolled in a clinical trial at a low-resource Gambian hospital were more likely to survive to hospital discharge than those receiving usual care. This survival advantage appeared to be most pronounced during the rainy season when baseline mortality was highest. These findings highlight that even severely unwell newborns can safely participate in clinical trials in resource-limited, high-mortality settings, offering reassurance to families, health workers, and ethical review boards. The participation benefit is likely attributable to higher quality of clinical care enabled by research-related strengthening of clinical monitoring, particularly due to staffing capacity, safety and governance oversight, and access to microbiological diagnostics. It is plausible that this benefit may be especially impactful during periods of high patient volume and health-system strain (Figure 2).

**Figure 2.**
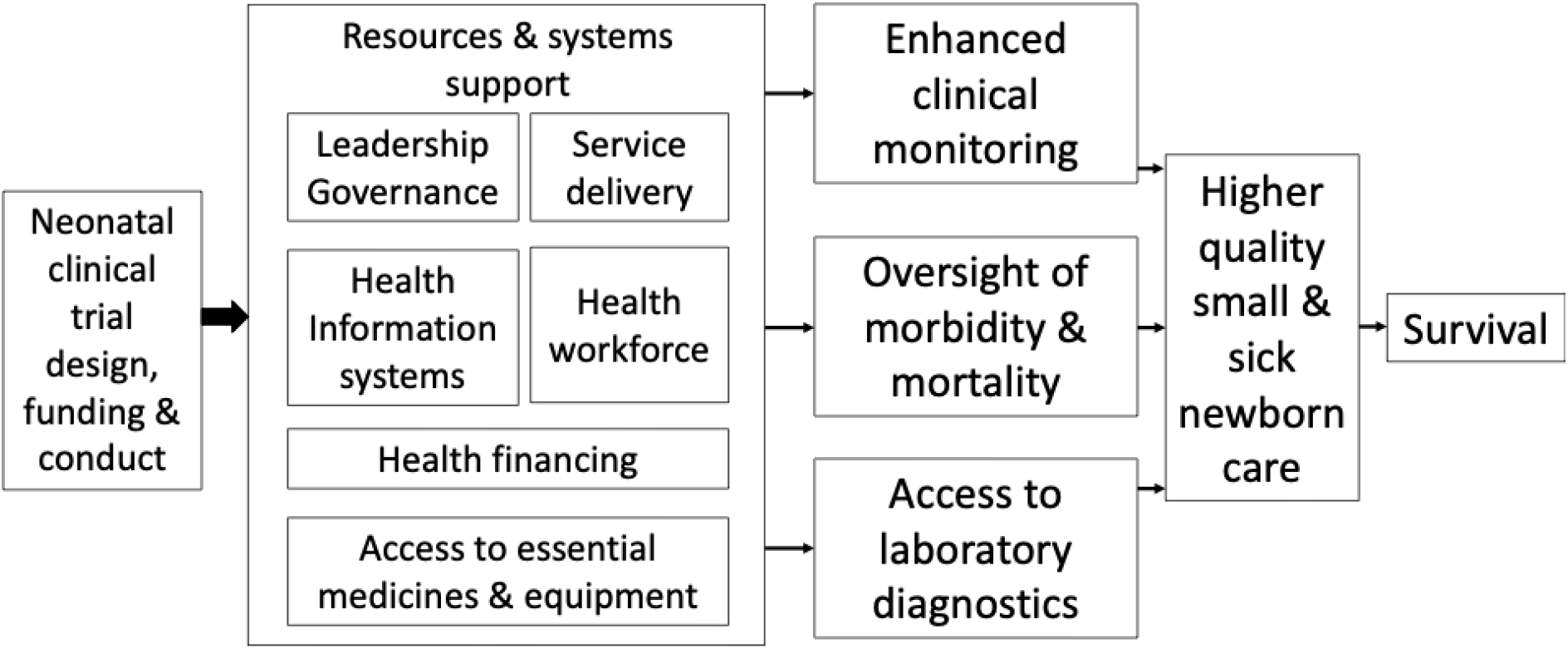
Direct acyclic graph for how research-related improvements to small and sick newborn care may improve neonatal survival in resource-limited health facility settings

To our knowledge, this is the first published evidence quantifying a neonatal trial participation survival benefit in a LMIC. Previous studies assessing trial participation effects were conducted in well-resourced hospitals in HIC and generally reported no impact on short- or long-term outcomes for preterm neonates [7, 8], aside from one small North American study that reported a reduction in ventilation duration [6]. Trial participation effects are highly likely to vary with health-system capacity and context. In settings where research resources mitigate health-system limitations and strain, participation effects are likely to be more pronounced. This interpretation is supported by our observation that the trial participation effect appeared to be stronger during the rainy season, when preterm/LBW admission rates [18], illness severity, and baseline mortality (Table 3) are highest. Accordingly, our findings may not be generalisable to better resourced LMIC facilities or contexts with lower baseline mortality. Our findings are also not relevant for observational studies using only routinely collected data without research-related care or systems improvements.

Most neonatal deaths are attributable to gaps in basic care [23], and an estimated 750,000 deaths could be prevented annually with scale-up of existing evidence-based interventions [19] and provision of high quality SSNC. Our finding of a 6% absolute and 29% relative survival benefit, driven by effects observed during the high-burden rainy season, highlights the extent to which accelerated survival gains for SVN are achievable through provision of higher quality SSNC when systems are under greatest strain. Health-service readiness assessments identified three research-related enhancements likely to have contributed to the observed mortality reduction: 1) Strengthened clinical monitoring, including continuous pulse oximetry and higher nursing to patient ratios; 2) Rigorous clinical oversight processes and strict adherence to WHO guidelines with prospective compliance checks; 3) Free access to essential laboratory diagnostics, particularly blood cultures.

Pulse oximetry provides real-time heart rate and oxygen saturation measurements, essential for guiding SSNC, reducing morbidity, and improving neonatal survival across all settings [24, 25]. During the eKMC trial, neonates receiving oxygen underwent continuous pulse oximetry monitoring in accordance with WHO guidance [26], whereas usual-care neonates received only intermittent spot checks due to limited device availability, maintenance challenges, and lack of reliable power sources. The eKMC trial provided additional pulse oximeters, neonatal-sized probes, re-chargeable batteries and systems to ensure availability of charged batteries, and maintenance support through the MRCG biomedical department. These enhancements are likely to have been particularly impactful during the rainy season, when higher admission rates and ward congestion may have increased the risk that clinical deterioration was undetected. This underscores the need for pulse oximeter provision to be accompanied by systems strengthening to ensure reliable and well-maintained medical device use, consistent with the NEST360 implementation framework [27].

The trained nurse to neonate ratio also differed markedly between groups, varying from minimum values of one nurse to one trial participant for eKMC participants to one nurse to 20 usual care neonates. Low nurse-to-patient ratios are associated with sub-optimal SSNC delivery in low-resource, high-mortality LMIC health-facilities [28], typically exacerbated during seasonal admission surges. During the rainy season, when nursing staffing levels remained fixed despite increased patient admissions, additional trained nurses with responsibility only for trial participants likely played a critical role in promoting compliance with high-priority tasks such as regular vital-sign assessment and timely detection of clinical deterioration [29]. Although research staff also undertook screening, enrolment, and data collection duties, most of their daily activities involved direct clinical monitoring and review of care plans and adverse event monitoring, increasing both the consistency and quality of SSNC at a time when routine services were most stretched. The trial’s safety and SAE review processes provided an additional oversight mechanism analogous to the Maternal and Perinatal Death Surveillance and Response System (MPDSR), supporting continuous SSNC quality improvement [23], which may be especially valuable during periods of heightened service pressure.

Laboratory diagnostics, particularly blood cultures, were freely available for trial participants through MRCG laboratories and were crucial given that 18% of trial participants developed suspected or confirmed late onset sepsis [14]. Most invasive bacterial isolates identified during eKMC trial were resistant to first-line antibiotics (73%, 8/11)[14], and infection accounted for 59% (37/63) of fatal SAEs. Despite on-site microbiology capacity at EFSTH, routine blood culture for usual-care neonates were not available due to the lack of Bactec bottles. This diagnostic gap aligns with findings from elsewhere in Africa that fewer than half of tertiary or secondary hospitals with microbiological capabilities in Kenya, Tanzania, Malawi, and Nigeria routinely perform neonatal blood cultures [30]. The trial site also experienced an outbreak of MDR *Klebsiella pneumoniae [31]* with high neonatal carriage of MDR *Klebsiella pneumoniae, E coli*, and *Acinetobacter* species [32] documented the year preceding eKMC trial onset. In such a high-bacterial AMR context, lack of blood cultures likely contributed substantially to poorer outcomes through ineffective empirical antibiotic treatment. Seasonal increases in bacterial infections during the rainy season are also well described in The Gambia [33], and it is possible that improved access to blood culture may have contributed to our finding of a seasonal participation effect.

Recruiting unwell neonates into clinical trials is challenging in all settings, particularly when evaluating time-critical interventions such as early KMC. Nevertheless, our finding of a substantial survival benefit, most evident during periods of highest baseline risk, provides important reassurance that trial participation is safe even for very sick newborns. The estimated 6% absolute mortality risk reduction is also valuable for researchers planning future SSNC intervention trials in similar settings, to inform more accurate sample size calculations and reduce the risk of under-powered studies.

This study has several limitations. First, our findings reflect participation effects in an individually randomised trial and may not apply to cluster-randomised trials or unit-level intervention designs. Similarly, the findings may not be generalisable to community-based studies or intervention trials not requiring enhanced clinical monitoring or safety oversight. Although we adjusted for known confounders such as twin status [34], other unmeasured clinical differences between groups may have influenced outcomes. The most common reason for non-enrolment in the usual-care group was caregiver unavailability, which may have been associated with severe maternal illness, a known confounding factor for neonatal morbidity. However, we consider this potential difference unlikely to be clinically meaningful, as both groups had comparable stability scores and were classified as mildly unstable at baseline. Lack of comprehensive data from non-enrolled infants prevented adjustment for possible differences in CPAP use, an important driver of neonatal survival [35]. We lacked data on KMC delivery among non-enrolled infants and cannot exclude the possibility that greater KMC exposure among enrolled infants contributed to the observed associations, despite no detectable mortality benefit within the eKMC trial. However, both trial arms received less than the minimum KMC duration previously associated with mortality benefit [15, 36], suggesting that any such effect was likely limited.

## CONCLUSION

Participation in an individually randomised clinical trial at a high-mortality, low resource Gambian hospital was associated with a substantial survival benefit for preterm/LBW newborns. This association appeared to be largely restricted to the rainy season, when mortality rates were highest, suggesting that benefits of trial participation may be greatest in higher-mortality contexts. The survival benefit was most likely driven by higher-quality SSNC enabled through optimised clinical monitoring, improved staffing availability, rigorous safety and governance oversight, and access to microbiological diagnostics. These components are foundational for improving neonatal survival and should be urgently prioritised within national and regional programmes to accelerate progress towards meeting SDG target 3.2 by 2030.

## Supporting information

Supplementary material (STROBE checklist, Table S1, Table S2)

## DECLARATIONS

### Data sharing statement

De-identified individual participant data are available upon request to the PI and in accordance with MRCG institutional regulations.

### Patient consent for publication

Not applicable

### Authors Contributions

HB and JEL conceived the research question, obtained the funding and designed the parent trial with input from AR, and SC. The trial was implemented by HB, AG, BK, GW and ALS. Data was collected and co-ordinated by AG, BK and GW. YN was responsible for data cleaning and verification. Service readiness data was collected by AG, GW, BK, SK and AH. UO was the trial safety monitor. ALS and MB facilitated implementation of the parent trial. HB performed the statistical analyses and wrote the original draft of the manuscript with critical revision for important intellectual content from SC, JEL, AR, UO. All authors approved the final manuscript as submitted. HB agrees to be accountable for all aspects of the work and takes responsibility for the decision to publish.

## Acknowledgements

We would like to thank all the newborns and families who engaged with the eKMC trial along with the EFSTH neonatal unit staff and management team, eKMC field team, and the Gambian Government Ministry of Health. An earlier version of this work was previously submitted to Global Health Network Conference 2022 (doi.10.21428/3d48c34a.dc3b6600).

