## Supplementary material (STROBE checklist, Table S1, Table S2) for "Short-term survival benefit associated with neonatal clinical trial participation: An observational cohort study in The Gambia"

### 1. STROBE checklist

Checklist of items that should be included in reports of ***cohort studies***

| | Item No | Recommendation | Page No |
|---|---|---|---|
| **Title and abstract** | 1 | (*a*) Indicate the study's design with a commonly used term in the title or the abstract | 1 |
| | | (*b*) Provide in the abstract an informative and balanced summary of what was done and what was found | 1-2 |
| **Introduction** | | | |
| Background/rationale | 2 | Explain the scientific background and rationale for the investigation being reported | 3-4 |
| Objectives | 3 | State specific objectives, including any prespecified hypotheses | 4 |
| **Methods** | | | |
| Study design | 4 | Present key elements of study design early in the paper | 4 |
| Setting | 5 | Describe the setting, locations, and relevant dates, including periods of recruitment, exposure, follow-up, and data collection | 4-5 |
| Participants | 6 | (*a*) Give the eligibility criteria, and the sources and methods of selection of participants. Describe methods of follow-up | 5 |
| | | (*b*) For matched studies, give matching criteria and number of exposed and unexposed | NA |
| Variables | 7 | Clearly define all outcomes, exposures, predictors, potential confounders, and effect modifiers. Give diagnostic criteria, if applicable | 5 |
| Data sources/ measurement | 8 | For each variable of interest, give sources of data and details of methods of assessment (measurement). Describe comparability of assessment methods if there is more than one group | 5 |
| Bias | 9 | Describe any efforts to address potential sources of bias | NA |
| Study size | 10 | Explain how the study size was arrived at | 5 |
| Quantitative variables | 11 | Explain how quantitative variables were handled in the analyses. If applicable, describe which groupings were chosen and why | 6 |
| Statistical methods | 12 | (*a*) Describe all statistical methods, including those used to control for confounding | 6 |
| | | (*b*) Describe any methods used to examine subgroups and interactions | 6 |
| | | (*c*) Explain how missing data were addressed | 6 |
| | | (*d*) If applicable, explain how loss to follow-up was addressed | NA |
| | | (*e*) Describe any sensitivity analyses | NA |
| **Results** | | | |
| Participants | 13 | (a) Report numbers of individuals at each stage of study—eg numbers potentially eligible, examined for eligibility, confirmed eligible, included in the study, completing follow-up, and analysed | 6 |
| | | (b) Give reasons for non-participation at each stage | 6 |
| | | (c) Consider use of a flow diagram | 16 |
| Descriptive data | 14 | (a) Give characteristics of study participants (eg demographic, clinical, social) and information on exposures and potential confounders | 6, 16 |
| | | (b) Indicate number of participants with missing data for each variable of interest | 16 |
| | | (c) Summarise follow-up time (eg, average and total amount) | NA |
| Outcome data | 15 | Report numbers of outcome events or summary measures over time | 7 |

| | | | |
|---|---|---|---|
| Main results | 16 | (*a*) Give unadjusted estimates and, if applicable, confounder-adjusted estimates and their precision (eg, 95% confidence interval). Make clear which confounders were adjusted for and why they were included | 7 |
| | | (*b*) Report category boundaries when continuous variables were categorized | 16 |
| | | (*c*) If relevant, consider translating estimates of relative risk into absolute risk for a meaningful time period | 7 |
| Other analyses | 17 | Report other analyses done—eg analyses of subgroups and interactions, and sensitivity analyses | 7 |
| **Discussion** | | | |
| Key results | 18 | Summarise key results with reference to study objectives | 7-9 |
| Limitations | 19 | Discuss limitations of the study, taking into account sources of potential bias or imprecision. Discuss both direction and magnitude of any potential bias | 9 |
| Interpretation | 20 | Give a cautious overall interpretation of results considering objectives, limitations, multiplicity of analyses, results from similar studies, and other relevant evidence | 7-9 |
| Generalisability | 21 | Discuss the generalisability (external validity) of the study results | 7 |
| **Other information** | | | |
| Funding | 22 | Give the source of funding and the role of the funders for the present study and, if applicable, for the original study on which the present article is based | 10 |

### 2. Additional tables

Supplementary Table S1. Reasons for non-enrolment to eKMC trial (n=266)

| **Reason for non-enrolment** | **Number (%) N= 266** |
|---|---|
| No caregiver available | 159 (59.8) |
| No study bed | 69 (25.9) |
| Consent declined | 31 (11.7) |
| Other[a] | 6 (2.3) |
| Not known[b] | 1 (0.4) |

a) Other reasons for non-enrolment included team training (n=5) and security issues preventing research staff from being present at the site (n=1); b) The one neonate for whom non enrolment reason was not known met all clinical eligibility criteria but the exact reason for non-enrolment was not recorded

Supplementary Table S2. Baseline characteristics for eligible neonates not enrolled, stratified by reason for non-enrolment

| | **No caregiver N=159** | **No study bed N=69** | **No consent N=31** |
|---|---|---|---|
| Male sex, Nº (%)[a] | 72 (45) | 29 (42) | 14 (45) |
| Weight (g), median (IQR) | 1544 (1197-1760) | 1400 (1100-1664) | 1550 (1332-1694) |
| Weight distribution, Nº (%) | | | |
| <1200g | 40 (25) | 20 (29) | 2 (6) |
| ≥1200g | 119 (75) | 49 (71) | 29 (94) |
| Twin, Nº (%) | 31 (19) | 18 (26) | 10 (32) |
| Admission age (h), median (IQR)[b] | 1.08 (0.67-3.35) | 1.17 (0.65-3.0) | 1.97 (0.63-4.5) |
| Admitted during night shift,[b] Nº (%) | 69 (45) | 28 (41) | 15 (48) |
| Admitted during rainy season, Nº (%) | 76 (48) | 44 (64) | 15 (48) |
| In-born (EFSTH),[c] Nº (%) | 105 (66) | 42 (63) | 13 (42) |
| Axillary temperature (°C), median (IQR) | 36.3 (35.5-37.0) | 36.1 (35.4-36.7) | 36.3 (35.8-36.9) |
| Respiratory rate (CPM), median (IQR) | 58 (48-70) | 56 (47-66) | 61 (51-67) |
| Heart rate (BPM), median (IQR) | 141 (131-155) | 138 (133-169) | 147 (141-158) |
| $SpO_2$ (%), median (IQR)[d] | 97.3 (95-98.4) | 96.9 (90.5-98.2) | 96.3 (94.5-98) |
| NMR-2000 score, median (IQR)[d] | 18.5 (14.9-20.5) | 18.1 (15.2-19.4) | 18.5 (16.6-19.9) |

BPM = beats per minute; CPM = Cycles per minute; EFSTH = Edward Francis Small Teaching Hospital; IQR = Interquartile range.

a) Missing data on sex for n=1 in no study bed group; b) Missing data on admission time for n=5 in no caregiver group and n=1 in no study bed group; c) Missing data on place of birth for n=2 in no study bed group; d) Missing data on SpO2 levels for n=14 in no caregiver group, n=13 in no study bed group, and n=3 in no consent group.
